# Using General Practice Patient Survey data to explore prevalence and patient uncertainty about Long Covid

**DOI:** 10.1101/2024.10.25.24316047

**Authors:** Mirembe Woodrow, Nida Ziauddeen, Dianna Smith, Nisreen A Alwan

## Abstract

**Background:** The high global burden of Long Covid (LC) has significant implications for population wellbeing, healthcare, social care and national economies.

**Aim:** To explore associations between patient sociodemographic and health characteristics with two outcomes: reporting having LC and expressing uncertainty about having LC, as described by general practice (GP) survey respondents.

**Design and setting:** Analysis of GP Patient Survey (England), a random sample of 759,149 patients aged 16yrs+ registered with a GP in England (2023).

**Method:** Multivariable logistic regression modelling comparing those with and without LC, and those who were unsure in relation to patient characteristics.

**Results:** 4.8% of respondents reported having LC, and 9.1% were unsure. Significant adjusted associations indicating higher risk of LC included age (highest odds 35-54yrs), sex (females), ethnicity (White Gypsy/Irish Traveller, mixed/multiple ethnic groups), sexual orientation (gay/lesbian or bisexual), living in a deprived area, being a carer or a parent and having a long-term condition (LTC). Those aged ≤25yrs, males, non-binary, heterosexual, not parents or carers, from other white, Indian, Bangladeshi, Chinese, Black or Arab backgrounds, former and current smokers, and with no defined LTC were more likely to be unsure about having LC compared to answering ‘yes’.

**Conclusion:** There is an unequal distribution of LC in England, with the condition being more prevalent in minoritised and disadvantaged groups. There are also high levels of uncertainty about having LC. Improved awareness is needed amongst the general population and healthcare professionals to ensure those most vulnerable in society are identified and provided with care and support.

## Introduction

In the UK, the Office for National Statistics estimated in March 2024 that 2 million adults and children experience self-reported Long Covid (LC) (3.3% of the population), with 69% and 41% experiencing it for at least 1 and 2 years respectively and 19% reporting that LC limits their ability to undertake day to day activities ‘a lot’^1^. There is also evidence of inequalities in LC prevalence, with higher prevalence associated with being female^2–9^, of older or middle age^2–4, 6, 7^, having a higher body mass index (BMI)^2, 3, 5, 7, 8^, smoking^3, 5^, and belonging to an ethnic minority group^5, 8^. Higher prevalence is also associated with greater deprivation^3–5, 9^. There is also unequal impact of the condition on people’s lives^10, 11^ ^12^ and inequitable support^13, 14^ ^15^ for the condition. More research is needed to understand the extent of this inequality and the action needed to address it.

As a relatively novel condition, knowledge about LC is still in its infancy amongst researchers and healthcare professionals but there is even more lack of awareness, exacerbated by barriers and stigma, in the general population leaving unwell people unsure if they have LC^16, 17^. This high and unequal burden of illness from LC has significant implications for society in terms of the infrastructure of accessible support and treatment needed for people with LC.

The General Practice Patient Survey (GPPS) is an annual survey of people aged 16yrs+ registered with a GP in England, administered by Ipsos on behalf of NHS England^18^. Established in 2007, it asks respondents about their experience of their local GP, other local NHS services and respondents’ general health. In 2022 a new question was added that asked whether respondents had Long Covid.

The aim of this study was to (1) explore prevalence of LC, (2) examine its potential sociodemographic and health characteristics and (3) explore factors associated with being unsure about having LC.

## Methods

### Sampling

Each year in January the GPPS is issued by post to a random sample of patients registered with a GP in England. It can be completed on paper (returned using a FREEPOST envelope), online or using a dedicated telephone helpline. Invitations and reminders are also issued by text message and the online survey is available in 14 different languages and British Sign Language. Paper copies can be requested in Braille and large print. Data is grouped by geographical and health system factors including general practice, Primary Care Network, Integrated Care System, Local Authority and NHS Region. Indices of deprivation are also available according to patient postcode and further technical detail is available online^19^.

The survey is issued to approximately 2.5 million registered patients and aimed to ensure a minimum number of responses per practice and Primary Care Network (PCN) and a set confidence interval for each practice. A proportionately stratified (by age, gender and postcode) unclustered sample is drawn for each practice.^19^ Patient identifiable data is not collected. It is a standalone self-reported survey that is not linked to GP patient records. The questionnaire was last significantly revised in 2021 and is available online^20^. More detail about the survey is available at https://gp-patient.co.uk/FAQ

### Design

This is an analysis of cross-sectional survey data using the GPPS dataset for 2023. Permission to access and use the individual patient level survey data from 2023 was requested and granted by agreement between NHS England and the University of Southampton. Ethical approval for this analysis was granted by the University of Southampton Faculty of Medicine Ethics Committee (No.82369).

### Variables

The outcome was the response to the question ‘Would you describe yourself as having “Long Covid”, that is, you are still experiencing symptoms more than 12 weeks after you first had COVID-19, that are not explained by something else?’. Respondents could select ‘yes’, ‘no’, ‘not sure’ or ‘prefer not to say’.

The exposure variables used in the analysis are listed in Tables 1 and 2. All were categorical and used as collected with the exception of smoking status. Smoking status was recorded as never, former, occasional and regular smoker; and was condensed to never, former and current smoker (regular and occasional) for analysis.

**Table 1:** Characteristics of the study population by Long Covid status as reported by survey participants (n=759,149)

|  | Yes | No | Not sure | Prefer not to say |
| --- | --- | --- | --- | --- |
|  | n (%) | n (%) | n (%) | n (%) |
| **All respondents** |  |  |  |  |
|  | 35445 (4.8) | 632748 (85.3) | 67257 (9.1) | 5991 (0.8) |
| **Commissioning region** |  |  |  |  |
| East | 3379 (4.3) | 68103 (86.7) | 6625 (8.4) | 486 (0.6) |
| London | 6158 (4.7) | 110489 (84.2) | 12859 (9.8) | 1732 (1.3) |
| Midlands | 7450 (5.1) | 124690 (84.6) | 13922 (9.5) | 1319 (0.9) |
| North East | 5687 (5.1) | 94826 (84.9) | 10408 (9.3) | 800 (0.7) |
| North West | 6095 (5.5) | 93015 (84.0) | 10781 (9.7) | 890 (0.8) |
| South East | 4111 (4.2) | 84642 (87.3) | 7728 (8.0) | 519 (0.5) |
| South West | 2565 (3.9) | 56983 (88.0) | 4934 (7.6) | 245 (0.4) |
| **Sex** |  |  |  |  |
| Female | 20973 (5.1) | 353888 (85.2) | 37635 (9.1) | 2649 (0.6) |
| Male | 13444 (4.4) | 263956 (86.1) | 27246 (8.9) | 2062 (0.7) |
| Non-binary | 102 (6.5) | 1214 (76.7) | 224 (14.2) | 42 (2.7) |
| Prefer to self-describe | 82 (8.2) | 693 (69.3) | 153 (15.3) | 72 (7.2) |
| Prefer not to say | 391 (4.9) | 5502 (69.4) | 1104 (13.9) | 937 (11.8) |
| **Age** |  |  |  |  |
| 16-24 years | 1148 (4.4) | 22183 (84.9) | 2391 (9.2) | 403 (1.5) |
| 25-34 years | 2759 (4.9) | 47858 (84.4) | 5327 (9.4) | 745 (1.3) |
| 35-44 years | 5039 (5.7) | 73225 (82.3) | 9535 (10.7) | 1171 (1.3) |
| 45-54 years | 7313 (6.2) | 95929 (81.8) | 12793 (10.9) | 1184 (1.0) |
| 55-64 years | 9027 (5.6) | 135435 (83.8) | 16107 (10.0) | 1079 (0.7) |
| 65-74 years | 6349 (4.1) | 137287 (87.5) | 12556 (8.0) | 684 (0.4) |
| 75-84 years | 2946 (2.9) | 92496 (90.3) | 6577 (6.4) | 443 (0.4) |
| 85+ years | 664 (2.5) | 24716 (91.2) | 1557 (5.8) | 162 (0.6) |
| **Ethnicity** |  |  |  |  |
| White - English, Welsh, Scottish, Northern Irish or British | 25968 (4.7) | 479699 (86.5) | 46529 (8.4) | 2260 (0.4) |
| White – Irish | 362 (4.7) | 6565 (86.0) | 678 (8.9) | 31 (0.4) |
| White - Gypsy or Irish Traveller | 42 (14.6) | 203 (70.5) | 36 (12.5) | 7 (2.4) |
| White - Roma | 33 (5.1) | 495 (76.3) | 87 (13.4) | 34 (5.2) |
| White - Any other White background | 2185 (5.4) | 33154 (81.4) | 4692 (11.5) | 683 (1.7) |
| Mixed or Multiple ethnic groups - White and Black Caribbean | 154 (5.7) | 2214 (82.6) | 279 (10.4) | 35 (1.3) |
| Mixed or Multiple ethnic groups - White and Black African | 87 (5.7) | 1240 (81.6) | 167 (11.0) | 25 (1.7) |
| Mixed or Multiple ethnic groups - White and Asian | 209 (6.4) | 2674 (81.6) | 348 (10.6) | 46 (1.4) |
| Mixed or Multiple ethnic groups - Any other Mixed or multiple ethnic background | 263 (6.8) | 3063 (78.8) | 503 (12.9) | 59 (1.5) |
| Asian or Asian British - Indian | 1319 (4.7) | 23244 (81.9) | 3227 (11.4) | 592 (2.1) |
| Asian or Asian British – Pakistani | 1111 (6.4) | 13483 (77.1) | 2341 (13.4) | 551 (3.2) |
| Asian or Asian British - Bangladeshi | 368 (6.2) | 4522 (76.6) | 851 (14.4) | 159 (2.7) |
| Asian or Asian British - Chinese | 233 (4.0) | 4814 (82.1) | 725 (12.4) | 90 (1.5) |
| Asian or Asian British - Any other Asian background | 707 (5.4) | 10503 (80.1) | 1616 (12.3) | 279 (2.1) |
| Black, Black British, Caribbean or African - Caribbean | 349 (4.3) | 7007 (85.9) | 704 (8.6) | 100 (1.2) |
| Black, Black British, Caribbean or African - African | 523 (2.9) | 16244 (89.2) | 1219 (6.7) | 220 (1.2) |
| Black, Black British, Caribbean or African - Any other Black, Black British, Caribbean or African background | 193 (4.22) | 3902 (85.4) | 418 (9.1) | 58 (1.2) |
| Other ethnic group - Arab | 191 (5.1) | 2982 (79.2) | 492 (13.1) | 102 (2.7) |
| Any other ethnic group | 707 (6.0) | 9106 (77.7) | 1465 (12.5) | 441 (3.8) |
| **Sexual orientation** |  |  |  |  |
| Heterosexual | 31093 (4.7) | 568277 (86.0) | 58132 (8.8) | 3206 (0.5) |
| Gay or lesbian | 723 (6.5) | 9267 (83.1) | 1119 (10.0) | 47 (0.4) |
| Bisexual | 609 (6.8) | 7310 (81.2) | 990 (11.0) | 93 (1.0) |
| Other | 500 (6.5) | 6164 (79.8) | 868 (11.2) | 197 (2.6) |
| I would prefer not to say | 1939 (4.9) | 30491 (77.1) | 4916 (12.4) | 2192 (5.5) |
| **Gender identity is the same as the sex registered at birth** |  |  |  |  |
| Yes | 34344 (4.8) | 627191 (85.6) | 64753 (9.0) | 4543 (0.6) |
| No | 267 (6.3) | 3289 (77.9) | 534 (12.6) | 134 (3.2) |
| Prefer not to say | 451 (5.3) | 5626 (66.5) | 1244 (14.7) | 1140 (13.5) |
| **Patient area of residence IMD quintile** |  |  |  |  |
| 1 (most deprived) | 8917 (6.0) | 121888 (81.4) | 16715 (11.2) | 2207 (1.5) |
| 2 | 7606 (5.1) | 125690 (84.0) | 14726 (9.8) | 1589 (1.1) |
| 3 | 7082 (4.7) | 130237 (85.8) | 13464 (8.9) | 1021 (0.7) |
| 4 | 6414 (4.3) | 129861 (87.2) | 12011 (8.1) | 717 (0.5) |
| 5 (least) | 5408 (3.8) | 124732 (88.5) | 10300 (7.3) | 452 (0.3) |
| **Work status** |  |  |  |  |
| In full-time paid work (30 hours or more each week) | 13777 (5.4) | 213406 (84.2) | 24409 (9.6) | 1891 (0.8) |
| In part-time paid work (under 30 hours each week) | 4958 (5.4) | 77225 (84.1) | 8955 (9.8) | 667 (0.7) |
| In full-time education at school, college or university | 657 (4.4) | 12649 (85.7) | 1270 (8.6) | 188 (1.3) |
| Unemployed | 1425 (5.7) | 19703 (79.4) | 3068 (12.4) | 609 (2.5) |
| Permanently sick or disabled | 3222 (9.3) | 25998 (75.4) | 4886 (14.2) | 389 (1.1) |
| Fully retired from work | 7742 (3.1) | 222069 (89.8) | 16725 (6.8) | 873 (0.4) |
| Looking after the family or home | 1527 (4.7) | 26989 (83.3) | 3472 (10.7) | 404 (1.3) |
| Doing something else | 1033 (4.9) | 17284 (81.2) | 2277 (10.7) | 689 (3.2) |
| **Carer** |  |  |  |  |
| No | 24598 (4.3) | 492447 (86.4) | 48681 (8.5) | 4502 (0.8) |
| 1-9hrs per week | 4693 (6.1) | 63947 (83.0) | 8121 (10.5) | 331 (0.4) |
| 10-19hrs per week | 1436 (7.2) | 16099 (80.6) | 2296 (11.5) | 150 (0.8) |
| 20-34hrs per week | 986 (7.5) | 10444 (79.0) | 1626 (12.3) | 157 (1.2) |
| 35-49hrs per week | 832 (7.4) | 8786 (78.1) | 1435 (12.8) | 196 (1.7) |
| 50hrs+ per week | 2107 (6.3) | 27570 (82.4) | 3461 (10.4) | 313 (0.9) |
| **Parent or guardian** |  |  |  |  |
| Yes | 7760 (5.6) | 114317 (82.9) | 14185 (10.3) | 1683 (1.2) |
| No | 27353 (4.6) | 513023 (85.9) | 52426 (8.8) | 4148 (0.7) |
| **Smoking status** |  |  |  |  |
| Never smoker | 18997 (4.5) | 364068 (86.3) | 35260 (8.4) | 3542 (0.8) |
| Former smoker | 11834 (5.2) | 192805 (84.7) | 21851 (9.6) | 1129 (0.5) |
| Occasional/regular smoker | 4302 (5.0) | 70511 (82.5) | 9506 (11.1) | 1154 (1.4) |
| **Deaf person who uses sign language** |  |  |  |  |
| Yes | 447 (14.6) | 2075 (67.7) | 389 (12.7) | 154 (5.0) |
| No | 34527 (4.7) | 623399 (85.5) | 65998 (9.1) | 5662 (0.8) |
| **Long term conditions** |  |  |  |  |
| Any long-term condition* | 26200 (5.8) | 378567 (83.9) | 44453 (9.9) | 1948 (0.4) |
| Any long-term condition excluding LC (derived variable) | 23443 (5.7) | 343728 (83.9) | 40632 (9.9) | 1690 (0.4) |
| Alzheimer's disease or other cause of dementia | 545 (9.2) | 4922 (82.8) | 444 (7.5) | 32 (0.5) |
| Arthritis or ongoing problem with back or joints | 10936 (6.4) | 139453 (82.1) | 18915 (11.1) | 622 (0.4) |
| Autism or autism spectrum condition | 528 (8.8) | 4742 (78.7) | 710 (11.8) | 43 (0.7) |
| Blindness or partial sight | 621 (5.3) | 9794 (84.1) | 1162 (10.0) | 64 (0.6) |
| Breathing condition such as asthma or COPD | 7757 (9.0) | 66590 (77.6) | 11132 (13.0) | 390 (0.5) |
| Cancer (diagnosis or treatment in the last 5 years) | 1253 (3.9) | 28620 (88.0) | 2543 (7.8) | 94 (0.3) |
| Deafness or hearing loss | 2981 (5.2) | 48069 (84.6) | 5605 (9.9) | 194 (0.3) |
| Diabetes | 4161 (5.7) | 61316 (83.6) | 7432 (10.1) | 405 (0.6) |
| Heart condition such as angina or atrial fibrillation | 3199 (5.8) | 45937 (83.8) | 5510 (10.1) | 196 (0.4) |
| High blood pressure | 8584 (5.4) | 135479 (84.4) | 15788 (9.8) | 617 (0.4) |
| Kidney or liver disease | 1211 (6.7) | 14477 (80.6) | 2159 (12.0) | 115 (0.6) |
| Learning disability | 625 (7.8) | 6283 (78.5) | 1004 (12.5) | 95 (1.2) |
| Mental health condition | 6332 (9.2) | 52790 (76.6) | 9472 (13.7) | 369 (0.5) |
| Neurological condition such as epilepsy | 922 (6.5) | 11724 (82.9) | 1442 (10.2) | 53 (0.4) |
| Stroke (which affects day-to-day life) | 478 (6.1) | 6455 (82.3) | 856 (10.9) | 51 (0.7) |
| Another long-term condition or disability | 7830 (7.8) | 80496 (80.6) | 11186 (11.2) | 402 (0.4) |
| *No long-term condition* | *7176 (3.1)* | *208447 (88.7)* | *17565 (7.5)* | *1711 (0.7)* |
| **Religion** |  |  |  |  |
| None | 10477 (4.9) | 184896 (86.1) | 18564 (8.6) | 868 (0.4) |
| Buddhist | 290 (5.9) | 3959 (81.1) | 545 (11.2) | 85 (1.7) |
| Christian | 18359 (4.5) | 354716 (86.6) | 34526 (8.4) | 2002 (0.5) |
| Hindu | 713 (4.5) | 12980 (82.6) | 1741 (11.1) | 281 (1.8) |
| Jewish | 190 (4.4) | 3793 (88.5) | 281 (6.6) | 23 (0.5) |
| Muslim | 2457 (5.8) | 33111 (78.5) | 5420 (12.9) | 1176 (2.8) |
| Sikh | 377 (5.2) | 5821 (80.5) | 826 (11.4) | 211 (2.9) |
| Other | 883 (7.8) | 8892 (78.4) | 1412 (12.5) | 151 (1.3) |
| Prefer not to say | 1301 (5.4) | 18609 (76.9) | 3232 (13.4) | 1050 (4.3) |

\* Presented as reported in the survey, but this may include the LC outcome

**Table 2:** Odds of reporting having Long Covid compared to not having Long Covid (yes vs no) using General Practice Patient Survey data (2023)

|  | Unadjusted |  |  |  | Adjusted |  |  |  |
| --- | --- | --- | --- | --- | --- | --- | --- | --- |
|  | Odds ratio | 95% CI lower | 95% CI upper | p | Odds ratio | 95% CI lower | 95% CI upper | p |
| **Age** |  |  |  |  |  |  |  |  |
| 16-24yrs | ref |  |  |  | ref |  |  |  |
| 25-34yrs | 1.114 | 1.040 | 1.196 | 0.003 | 1.071 | 0.995 | 1.153 | 0.068 |
| 35-44yrs | 1.330 | 1.245 | 1.420 | <0.0001 | 1.216 | 1.133 | 1.306 | <0.0001 |
| 45-54yrs | 1.473 | 1.382 | 1.570 | <0.0001 | 1.232 | 1.151 | 1.318 | <0.0001 |
| 55-64yrs | 1.288 | 1.209 | 1.372 | <0.0001 | 1.008 | 0.942 | 1.078 | 0.819 |
| 65-74yrs | 0.894 | 0.838 | 0.953 | 0.001 | 0.662 | 0.617 | 0.709 | <0.0001 |
| 75-84yrs | 0.615 | 0.574 | 0.660 | <0.0001 | 0.429 | 0.398 | 0.463 | <0.0001 |
| 85+yrs | 0.519 | 0.471 | 0.572 | <0.0001 | 0.339 | 0.304 | 0.378 | <0.0001 |
| **Sex** |  |  |  |  |  |  |  |  |
| Female | ref |  |  |  | ref |  |  |  |
| Male | 0.859 | 0.841 | 0.879 | <0.0001 | 0.879 | 0.859 | 0.900 | <0.0001 |
| Non-binary | 1.418 | 1.15 | 1.736 | 0.001 | 0.904 | 0.722 | 1.130 | 0.375 |
| Prefer to self-describe | 1.997 | 1.587 | 2.511 | <0.0001 | 1.451 | 1.134 | 1.856 | 0.003 |
| Prefer not to say | 1.199 | 1.081 | 1.330 | 0.001 | 0.998 | 0.889 | 1.121 | 0.974 |
| **Sexual orientation** |  |  |  |  |  |  |  |  |
| Heterosexual | ref |  |  |  | ref |  |  |  |
| Gay or lesbian | 1.426 | 1.320 | 1.539 | <0.0001 | 1.203 | 1.110 | 1.303 | <0.0001 |
| Bisexual | 1.523 | 1.401 | 1.655 | <0.0001 | 1.193 | 1.092 | 1.303 | <0.0001 |
| Other | 1.483 | 1.352 | 1.625 | <0.0001 | 1.212 | 1.096 | 1.339 | <0.0001 |
| I would prefer not to say | 1.162 | 1.109 | 1.219 | <0.0001 | 0.990 | 0.937 | 1.046 | 0.714 |
| **Gender identity is the same as the sex registered at birth** |  |  |  |  |  |  |  |  |
| Yes | ref |  |  |  | NA |  |  |  |
| No | 1.459 | 1.287 | 1.653 | <0.0001 |  |  |  |  |
| Prefer not to say | 1.441 | 1.308 | 1.587 | <0.0001 |  |  |  |  |
| **Parent** |  |  |  |  |  |  |  |  |
| No | ref |  |  |  | ref |  |  |  |
| Yes | 1.273 | 1.240 | 1.307 | <0.0001 | 1.058 | 1.024 | 1.093 | 0.001 |
| **Carer** |  |  |  |  |  |  |  |  |
| No | ref |  |  |  |  |  |  |  |
| 1-9hrs per week | 1.469 | 1.423, | 1.517 | <0.0001 | 1.387 | 1.341 | 1.434 | <0.0001 |
| 10-19hrs | 1.786 | 1.689 | 1.888 | <0.0001 | 1.612 | 1.522 | 1.707 | <0.0001 |
| 20-34hrs | 1.890 | 1.768 | 2.020 | <0.0001 | 1.651 | 1.540 | 1.769 | <0.0001 |
| 35-49hrs | 1.896 | 1.764 | 2.038 | <0.0001 | 1.538 | 1.426 | 1.658 | <0.0001 |
| 50hrs+ | 1.530 | 1.461 | 1.602 | <0.0001 | 1.434 | 1.367 | 1.505 | <0.0001 |
| **Ethnicity** |  |  |  |  |  |  |  |  |
| White - English, Welsh, Scottish, Northern Irish or British | ref |  |  |  | ref |  |  |  |
| White – Irish | 1.019 | 0.916 | 1.133 | 0.735 | 1.060 | 0.949 | 1.183 | 0.302 |
|  | Odds ratio | 95% CI lower | 95% CI upper | p | Odds ratio | 95% CI lower | 95% CI upper | p |
| White - Gypsy or Irish Traveller | 3.822 | 2.741 | 5.329 | <0.0001 | 2.452 | 1.697 | 3.544 | <0.0001 |
| White - Roma | 1.232 | 0.866 | 1.752 | 0.247 | 1.024 | 0.701 | 1.495 | 0.903 |
| White - Any other White background | 1.217 | 1.164 | 1.274 | <0.0001 | 1.096 | 1.044 | 1.150 | <0.0001 |
| Mixed or Multiple ethnic groups - White and Black Caribbean | 1.285 | 1.091 | 1.514 | 0.003 | 1.033 | 0.872 | 1.222 | 0.708 |
| Mixed or Multiple ethnic groups - White and Black African | 1.296 | 1.042 | 1.611 | 0.020 | 1.031 | 0.820 | 1.297 | 0.791 |
| Mixed or Multiple ethnic groups - White and Asian | 1.444 | 1.254 | 1.663 | <0.0001 | 1.278 | 1.104 | 1.481 | 0.001 |
| Mixed or Multiple ethnic groups - Any other Mixed or multiple ethnic background | 1.586 | 1.398 | 1.800 | <0.0001 | 1.274 | 1.116 | 1.454 | <0.0001 |
| Asian or Asian British - Indian | 1.048 | 0.990 | 1.110 | 0.104 | 0.939 | 0.855 | 1.031 | 0.185 |
| Asian or Asian British – Pakistani | 1.522 | 1.430 | 1.620 | <0.0001 | 1.077 | 0.975 | 1.189 | 0.146 |
| Asian or Asian British - Bangladeshi | 1.503 | 1.350 | 1.673 | <0.0001 | 1.037 | 0.907 | 1.186 | 0.590 |
| Asian or Asian British – Chinese | 0.894 | 0.783 | 1.020 | 0.097 | 0.864 | 0.751 | 0.994 | 0.041 |
| Asian or Asian British - Any other Asian background | 1.243 | 1.151 | 1.343 | <0.0001 | 1.061 | 0.971 | 1.160 | 0.189 |
| Black, Black British, Caribbean or African - Caribbean | 0.920 | 0.826 | 1.025 | 0.131 | 0.758 | 0.677 | 0.849 | <0.0001 |
| Black, Black British, Caribbean or African – African | 0.595 | 0.545 | 0.649 | <0.0001 | 0.470 | 0.428 | 0.517 | <0.0001 |
|  | Odds ratio | 95% CI lower | 95% CI upper | p | Odds ratio | 95% CI lower | 95% CI upper | p |
| Black, Black British, Caribbean or African - Any other Black, Black British, Caribbean or African background | 0.914 | 0.790 | 1.056 | 0.223 | 0.748 | 0.644 | 0.870 | <0.0001 |
| Other ethnic group - Arab | 1.183 | 1.022 | 1.370 | 0.025 | 0.879 | 0.743 | 1.041 | 0.136 |
| Any other ethnic group | 1.434 | 1.327 | 1.530 | <0.0001 | 1.154 | 1.057 | 1.260 | 0.001 |
| **Patient IMD quintile** |  |  |  |  |  |  |  |  |
| 1 (most deprived) | 1.687 | 1.630 | 1.747 | <0.0001 | 1.470 | 1.416 | 1.526 | <0.0001 |
| 2 | 1.396 | 1.347 | 1.446 | <0.0001 | 1.283 | 1.236 | 1.333 | <0.0001 |
| 3 | 1.254 | 1.210 | 1.300 | <0.0001 | 1.198 | 1.154 | 1.244 | <0.0001 |
| 4 | 1.139 | 1.098 | 1.182 | <0.0001 | 1.106 | 1.065 | 1.150 | <0.0001 |
| 5 (least) | ref |  |  |  |  |  |  |  |
| **Work status** |  |  |  |  |  |  |  |  |
| In full-time paid work (30 hours or more each week) | ref |  |  |  | NA |  |  |  |
| In part-time paid work (under 30 hours each week) | 0.994 | 0.962 | 1.028 | 0.746 |  |  |  |  |
| In full-time education at school, college or university | 0.805 | 0.742 | 0.872 | <0.0001 |  |  |  |  |
| Unemployed | 1.120 | 1.059 | 1.185 | <0.0001 |  |  |  |  |
| Permanently sick or disabled | 1.920 | 1.844 | 2.000 | <0.0001 |  |  |  |  |
| Fully retired from work | 0.540 | 0.525 | 0.556 | <0.0001 |  |  |  |  |
| Looking after the family or home | 0.876 | 0.830 | 0.925 | <0.0001 |  |  |  |  |
| Doing something else | 0.926 | 0.867 | 0.988 | 0.020 |  |  |  |  |
| **Religion** |  |  |  |  |  |  |  |  |
| None | ref |  |  |  | ref |  |  |  |
| Buddhist | 1.293 | 1.146 | 1.459 | 0.001 | 1.260 | 1.104 | 1.438 | 0.001 |
| Christian | 0.913 | 0.891 | 0.936 | <0.0001 | 1.051 | 1.024 | 1.080 | <0.0001 |
| Hindu | 0.969 | 0.897 | 1.048 | 0.434 | 1.091 | 0.975 | 1.221 | 0.128 |
| Jewish | 0.884 | 0.763 | 1.024 | 0.100 | 0.980 | 0.836 | 1.149 | 0.805 |
| Muslim | 1.310 | 1.251 | 1.370 | <0.0001 | 1.191 | 1.099 | 1.291 | <0.0001 |
| Sikh | 1.143 | 1.028 | 1.271 | 0.013 | 1.279 | 1.110 | 1.474 | 0.001 |
|  | Odds ratio | 95% CI lower | 95% CI upper | p | Odds ratio | 95% CI lower | 95% CI upper | p |
| Other | 1.752 | 1.631 | 1.883 | <0.0001 | 1.538 | 1.425 | 1.659 | <0.0001 |
| Prefer not to say | 1.234 | 1.162 | 1.310 | <0.0001 | 1.246 | 1.166 | 1.332 | <0.0001 |
| **Smoking** |  |  |  |  |  |  |  |  |
| Never smoked | ref |  |  |  | ref |  |  |  |
| Former smoker | 1.176 | 1.149 | 1.204 | <0.0001 | 1.190 | 1.160 | 1.221 | <0.0001 |
| Occasional or regular smoker | 1.169 | 1.130 | 1.210 | <0.0001 | 0.959 | 0.925 | 0.995 | 0.024 |
| **Deaf person who uses sign language** |  |  |  |  |  |  |  |  |
| No | ref |  |  |  | NA |  |  |  |
| Yes | 3.890 | 3.510 | 4.311 | <0.0001 |  |  |  |  |
| **At least one Long Term Condition (excluding LC)** |  |  |  |  |  |  |  |  |
| No | ref |  |  |  | ref |  |  |  |
| Yes | 1.642 | 1.606 | 1.680 | <0.0001 | 1.965 | 1.916 | 2.014 | <0.0001 |

### Analysis

Data was analysed using Stata v17^21^. Descriptive statistics, frequencies and proportions were calculated followed by univariable and multivariable logistic regression. The primary analysis compared those answering ‘yes’ and ‘no’ to the LC question, excluding those answering unsure or ‘prefer not to say’. The same analysis comparing those answering ‘not sure’ and ‘no’ was also undertaken, as well as comparing ‘not sure’ and ‘yes’. We conducted the latter analysis to understand the factors associated with uncertainty about having the health condition which may act as a barrier to seeking care and support.

We conducted multivariable analysis to assess the independent effects of variables given they can potentially confound each other. The multivariable analysis included age, sex, sexuality, parental/guardian status, carer status, ethnicity, patient’s area of residence index of multiple deprivation (IMD) quintile (a combined index of relative deprivation based on data in seven domains)^22^, religion, smoking status and having at least one defined LTC (other than LC). A new variable was created to identify respondents with at least one defined LTC. The question that asked if respondents had any LTCs could not be included as respondents with LC could have counted their LC as a LTC (the LTC variable could include the LC outcome). Work status was included as an adjustment in the analysis comparing ‘yes’ and ‘not sure’ but not included when comparing ‘yes’ and ‘no’ (work status could have been impacted by having LC).

## Results

The survey was completed by 759,149 people (28.6% response rate).

Figure 1 shows the percentage of respondents self-reporting having LC and being unsure if they have LC.

**Figure 1:**
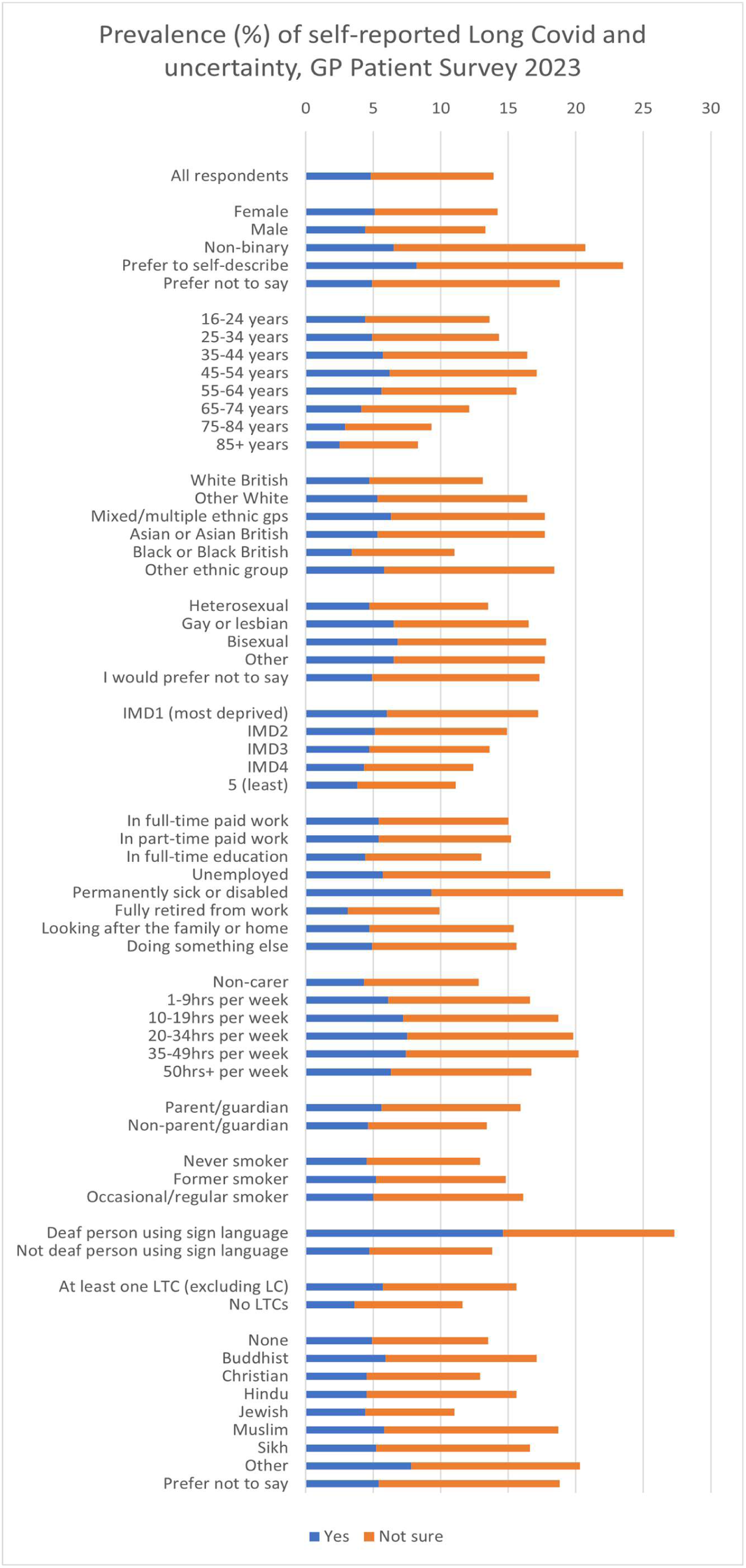
Prevalence of self-reported Long Covid and uncertainty, GP Patient Survey 2023.

### Respondents describing themselves as having LC (compared with those who answered ‘No’)

The percentage of respondents self-describing as having LC was 4.8% (n=35,445) (Table 1). The highest prevalence of LC was in the North West region (5.5%) and lowest in the South West (3.9%) (Table 1). 9% of people who described themselves as permanently sick or disabled had LC (Table 1). Compared to being in full-time work, people who were students, retired, looking after the family or home, or ‘doing something else’ were less likely to report having LC (Table 2). People who were unemployed or permanently sick or disabled were more likely to report having LC (unadjusted 1.12, 95%CI 1.06-1.19; 1.92, 95%CI 1.84-2.00 respectively).

Of those who stated they had LC, 59.9% (n=20,973) were female. Males were less likely to report having LC compared to females (adjusted OR 0.88, 95%CI 0.86-0.90) (Table 2). 60.7% of those with LC were aged between 35 and 64yrs. Compared to those aged 16-24yrs, having LC was significantly less common in those ≥65yrs and more common in those aged 25-64yrs even after adjustment for other characteristics (Table 2). The prevalence of LC significantly increased as area deprivation increased, with living in the most deprived area quintile associated with 47% higher odds compared to the least (adjusted OR 1.47, 95%CI 1.42-1.53) (Table 2).

People who were gay/lesbian, bisexual or ‘other’ sexual orientations had significantly higher prevalence compared to heterosexuals (Table 2). Parents were more likely to have LC compared to non-parents, as were carers in all caring hours categories compared to non-carers (Table 2). Compared to white British ethnicity, significantly higher adjusted odds of having LC were seen in White Gypsy or Irish Traveller groups, any other White background, White and Asian and ‘other’ mixed or multiple ethnic groups, and ‘any other ethnic group’ (Table 1). All three Black as well as Chinese ethnic groups were less likely to report having LC (Table 2). Compared to having no religion, adjusted ORs of having LC were higher for Buddhist, Christian, Muslim, Sikh and ‘other’ religions (Table 2).

The prevalence of LC amongst current regular smokers was 4.8% (Table 1) and the proportion of people with LC who were regular or occasional smokers was 12.2% compared to 11.2% smokers among those with no LC. Compared to people who had never smoked, the adjusted OR of having LC in occasional/regular smokers was 0.96 (95%CI 0.93-1.00) and for former smokers was 1.19 (95%CI 1.16-1.22) (Table 2).

There was particularly high prevalence of LC amongst people who were deaf and using sign language (14.6%), and those who had Alzheimer’s/dementia (9.2%), autism (8.8%), a breathing condition (9.0%), a mental health condition (9.2%) or another LTC not listed (7.8%) (Table 1). People with at least one defined LTC (which was not LC) had higher odds of having LC (adjusted OR 1.97, 95%CI 1.92-2.01).

### Respondents who were ‘not sure’ if they would describe themselves as having LC (compared with those who answered ‘Yes’)

Those aged 25-74yrs were significantly less likely to say they are unsure about having LC (compared to those who answered ‘yes’ to having LC) than those aged 16-24yrs (Table 3). Males and non-binary people were significantly more likely to be unsure compared to females. Gay/lesbian and ‘Other’ sexual orientations were significantly less likely to be unsure compared to heterosexuals. Parents were less likely to be unsure compared to non-parents (adjusted OR 0.93, 95% CI 0.89-0.96), and carers less likely to be unsure compared to non-carers (Table 3).

**Table 3:** Odds of being uncertain about having Long Covid compared to reporting having Long Covid (not sure vs yes) using General Practice Patient Survey data (2023)

|  | Unadjusted |  |  |  | Adjusted |  |  |  |
| --- | --- | --- | --- | --- | --- | --- | --- | --- |
|  | Odds ratio | 95% CI lower | 95% CI upper | p | Odds ratio | 95% CI lower | 95% CI upper | p |
| **Age** |  |  |  |  |  |  |  |  |
| 16-24yrs | ref |  |  |  | ref |  |  |  |
| 25-34yrs | 0.927 | 0.852 | 1.008 | 0.077 | 0.876 | 0.793 | 0.967 | 0.009 |
| 35-44yrs | 0.909 | 0.840 | 0.982 | 0.016 | 0.868 | 0.788 | 0.957 | 0.005 |
| 45-54yrs | 0.840 | 0.778 | 0.906 | <0.0001 | 0.850 | 0.774 | 0.935 | 0.001 |
| 55-64yrs | 0.857 | 0.795 | 0.923 | <0.0001 | 0.884 | 0.805 | 0.971 | 0.010 |
| 65-74yrs | 0.950 | 0.880 | 1.025 | 0.185 | 0.882 | 0.796 | 0.977 | 0.016 |
| 75-84yrs | 1.072 | 0.987 | 1.164 | 0.100 | 0.999 | 0.892 | 1.118 | 0.981 |
| 85+yrs | 1.126 | 1.004 | 1.263 | 0.043 | 1.097 | 0.947 | 1.271 | 0.216 |
| **Sex** |  |  |  |  |  |  |  |  |
| Female | ref |  |  |  | ref |  |  |  |
| Male | 1.129 | 1.100 | 1.160 | <0.0001 | 1.134 | 1.101 | 1.167 | <0.0001 |
| Non-binary | 1.224 | 0.968 | 1.548 | 0.092 | 1.394 | 1.077 | 1.805 | 0.012 |
| Prefer to self-describe | 1.040 | 0.795 | 1.360 | 0.776 | 1.035 | 0.775 | 1.382 | 0.815 |
| Prefer not to say | 1.573 | 1.400 | 1.768 | <0.0001 | 1.285 | 1.126 | 1.466 | <0.0001 |
| **Sexual orientation** |  |  |  |  |  |  |  |  |
| Heterosexual | ref |  |  |  | ref |  |  |  |
| Gay or lesbian | 0.828 | 0.753 | 0.910 | <0.0001 | 0.854 | 0.773 | 0.943 | 0.002 |
| Bisexual | 0.869 | 0.785 | 0.963 | 0.007 | 0.892 | 0.801 | 0.995 | 0.040 |
| Other | 0.929 | 0.831 | 1.037 | 0.190 | 0.877 | 0.776 | 0.991 | 0.035 |
| I would prefer not to say | 1.356 | 1.284 | 1.432 | <0.0001 | 1.186 | 1.113 | 1.264 | <0.0001 |
| **Gender identity is the same as the sex registered at birth** |  |  |  |  |  |  |  |  |
| Yes | ref |  |  |  | NA |  |  |  |
| No | 1.061 | 0.915 | 1.229 | 0.433 |  |  |  |  |
| Prefer not to say | 1.463 | 1.313 | 1.631 | <0.0001 |  |  |  |  |
| **Parent** |  |  |  |  |  |  |  |  |
| No | ref |  |  |  | ref |  |  |  |
| Yes | 0.954 | 0.924 | 0.984 | 0.003 | 0.926 | 0.890 | 0.964 | <0.0001 |
| **Carer** |  |  |  |  |  |  |  |  |
| No | ref |  |  |  | ref |  |  |  |
| 1-9hrs per week | 0.874 | 0.841 | 0.909 | <0.0001 | 0.916 | 0.879 | 0.954 | <0.0001 |
| 10-19hrs | 0.808 | 0.755 | 0.864 | <0.0001 | 0.822 | 0.766 | 0.883 | <0.0001 |
| 20-34hrs | 0.833 | 0.769 | 0.903 | <0.0001 | 0.829 | 0.761 | 0.902 | <0.0001 |
| 35-49hrs | 0.872 | 0.799 | 0.951 | 0.002 | 0.885 | 0.807 | 0.971 | <0.0001 |
| 50hrs+ | 0.830 | 0.785 | 0.878 | <0.0001 | 0.823 | 0.775 | 0.875 | <0.0001 |
| **Ethnicity** |  |  |  |  |  |  |  |  |
| White - English, Welsh, Scottish, Northern Irish or British | ref |  |  |  | ref |  |  |  |
| White – Irish | 1.045 | 0.919 | 1.189 | 0.499 | 0.993 | 0.867 | 1.137 | 0.919 |
|  | Odds ratio | 95% CI lower | 95% CI upper | p | Odds ratio | 95% CI lower | 95% CI upper | p |
| White - Gypsy or Irish Traveller | 0.478 | 0.306 | 0.747 | 0.001 | 0.508 | 0.310 | 0.831 | 0.007 |
| White - Roma | 1.471 | 0.985 | 2.197 | 0.059 | 1.354 | 0.880 | 2.082 | 0.168 |
| White - Any other White background | 1.198 | 1.137 | 1.264 | <0.0001 | 1.195 | 1.128 | 1.265 | <0.0001 |
| Mixed or Multiple ethnic groups - White and Black Caribbean | 1.011 | 0.830 | 1.232 | 0.913 | 1.025 | 0.836 | 1.258 | 0.810 |
| Mixed or Multiple ethnic groups - White and Black African | 1.071 | 0.826 | 1.389 | 0.603 | 1.083 | 0.823 | 1.426 | 0.568 |
| Mixed or Multiple ethnic groups - White and Asian | 0.929 | 0.782 | 1.104 | 0.404 | 0.884 | 0.737 | 1.061 | 0.186 |
| Mixed or Multiple ethnic groups - Any other Mixed or multiple ethnic background | 1.067 | 0.919 | 1.240 | 0.394 | 1.102 | 0.941 | 1.291 | 0.226 |
| Asian or Asian British - Indian | 1.365 | 1.278 | 1.458 | <0.0001 | 1.304 | 1.169 | 1.454 | <0.0001 |
| Asian or Asian British – Pakistani | 1.176 | 1.093 | 1.265 | <0.0001 | 1.118 | 0.995 | 1.256 | 0.061 |
| Asian or Asian British - Bangladeshi | 1.291 | 1.141 | 1.460 | <0.0001 | 1.224 | 1.048 | 1.430 | 0.011 |
| Asian or Asian British – Chinese | 1.737 | 1.497 | 2.014 | <0.0001 | 1.752 | 1.495 | 2.053 | <0.0001 |
| Asian or Asian British - Any other Asian background | 1.276 | 1.166 | 1.395 | <0.0001 | 1.266 | 1.140 | 1.405 | <0.0001 |
| Black, Black British, Caribbean or African - Caribbean | 1.126 | 0.989 | 1.281 | 0.072 | 1.169 | 1.019 | 1.340 | 0.026 |
| Black, Black British, Caribbean or African – African | 1.301 | 1.173 | 1.443 | <0.0001 | 1.351 | 1.207 | 1.513 | <0.0001 |
|  | Odds ratio | 95% CI lower | 95% CI upper | p | Odds ratio | 95% CI lower | 95% CI upper | p |
| Black, Black British, Caribbean or African - Any other Black, Black British, Caribbean or African background | 1.209 | 1.019 | 1.435 | 0.030 | 1.172 | 0.979 | 1.404 | 0.084 |
| Other ethnic group - Arab | 1.438 | 1.216 | 1.700 | <0.0001 | 1.461 | 1.203 | 1.775 | <0.0001 |
| Any other ethnic group | 1.156 | 1.056 | 1.267 | 0.002 | 1.126 | 1.014 | 1.250 | 0.026 |
| **Patient IMD quintile** |  |  |  |  |  |  |  |  |
| 1 (most deprived) | 0.984 | 0.944 | 1.026 | 0.455 | 0.971 | 0.927 | 1.016 | 0.200 |
| 2 | 1.017 | 0.974 | 1.061 | 0.454 | 0.993 | 0.948 | 1.040 | 0.765 |
| 3 | 0.998 | 0.956 | 1.043 | 0.936 | 0.989 | 0.945 | 1.036 | 0.642 |
| 4 | 0.983 | 0.940 | 1.028 | 0.458 | 0.987 | 0.942 | 1.035 | 0.586 |
| 5 (least) | ref |  |  |  | ref |  |  |  |
| **Work status** |  |  |  |  |  |  |  |  |
| In full-time paid work (30 hours or more each week) | ref |  |  |  | ref |  |  |  |
| In part-time paid work (under 30 hours each week) | 1.019 | 0.979 | 1.062 | 0.351 | 1.085 | 1.039 | 1.133 | <0.0001 |
| In full-time education at school, college or university | 1.091 | 0.991 | 1.202 | 0.077 | 0.917 | 0.816 | 1.031 | 0.146 |
| Unemployed | 1.215 | 1.137 | 1.298 | <0.0001 | 1.181 | 1.100 | 1.268 | <0.0001 |
| Permanently sick or disabled | 0.856 | 0.815 | 0.899 | <0.0001 | 0.937 | 0.887 | 0.990 | 0.020 |
| Fully retired from work | 1.219 | 1.178 | 1.262 | <0.0001 | 1.281 | 1.213 | 1.354 | <0.0001 |
| Looking after the family or home | 1.283 | 1.204 | 1.368 | <0.0001 | 1.391 | 1.297 | 1.491 | <0.0001 |
| Doing something else | 1.244 | 1.153 | 1.343 | <0.0001 | 1.243 | 1.146 | 1.348 | <0.0001 |
| **Religion** |  |  |  |  |  |  |  |  |
| None | ref |  |  |  | ref |  |  |  |
| Buddhist | 1.061 | 0.918 | 1.225 | 0.425 | 0.919 | 0.783 | 1.079 | 0.301 |
| Christian | 1.061 | 1.030 | 1.094 | <0.0001 | 1.023 | 0.989 | 1.057 | 0.185 |
| Hindu | 1.378 | 1.259 | 1.508 | <0.0001 | 1.082 | 0.949 | 1.234 | 0.238 |
| Jewish | 0.835 | 0.693 | 1.005 | 0.056 | 0.832 | 0.680 | 1.017 | 0.072 |
| Muslim | 1.245 | 1.180 | 1.313 | <0.0001 | 1.036 | 0.943 | 1.139 | 0.461 |
| Sikh | 1.237 | 1.092 | 1.400 | 0.001 | 0.968 | 0.820 | 1.144 | 0.706 |
|  | Odds ratio | 95% CI lower | 95% CI upper | p | Odds ratio | 95% CI lower | 95% CI upper | p |
| Other | 0.902 | 0.827 | 0.985 | 0.021 | 0.890 | 0.811 | 0.978 | 0.015 |
| Prefer not to say | 1.402 | 1.309 | 1.502 | <0.0001 | 1.215 | 1.125 | 1.313 | <0.0001 |
| **Smoking** |  |  |  |  |  |  |  |  |
| Never smoked | ref |  |  |  | ref |  |  |  |
| Former smoker | 0.995 | 0.967 | 1.024 | 0.721 | 1.053 | 1.020 | 1.086 | 0.001 |
| Occasional or regular smoker | 1.191 | 1.144 | 1.239 | <0.0001 | 1.274 | 1.220 | 1.330 | <0.0001 |
| **Deaf person who uses sign language** |  |  |  |  |  |  |  |  |
| Yes | ref |  |  |  | NA |  |  |  |
| No | 2.196 | 1.916 | 2.518 | <0.0001 |  |  |  |  |
| **At least one Long Term Condition (excluding LC)** |  |  |  |  |  |  |  |  |
| No | ref |  |  |  | ref |  |  |  |
| Yes | 0.781 | 0.761 | 0.803 | <0.0001 | 0.764 | 0.741 | 0.788 | <0.0001 |

White Gypsy or Irish Traveller were significantly less likely to be unsure that they had LC (compared to those who answered ‘yes’ to having LC) than White British ethnicity. ‘Any other White background’, Indian, Bangladeshi, Chinese, ‘Any other Asian background’, Black Caribbean, Black African, Arab and ‘Any other ethnic group’ were significantly more likely to be unsure compared to White British. There was no significant difference by level of area deprivation (IMD quintile) between those who were unsure and those who said they have LC or between those with a religion and those without.

Unemployment, working part-time, being retired, looking after the family or home, or ‘doing something else’ were positively linked to not being sure as opposed to saying ‘yes’ to have LC, compared with people working full-time. People who were permanently sick or disabled were less likely to be unsure (Table 3).

Compared with those who answered ‘yes’ to having LC, former or current smokers were significantly more likely to be unsure than people who never smoked. People with at least one defined LTC were less likely to be unsure compared to people without (Table 3).

Results comparing responses of those answering ‘not sure’ and ‘no’ are included in Table 4.

**Table 4:**
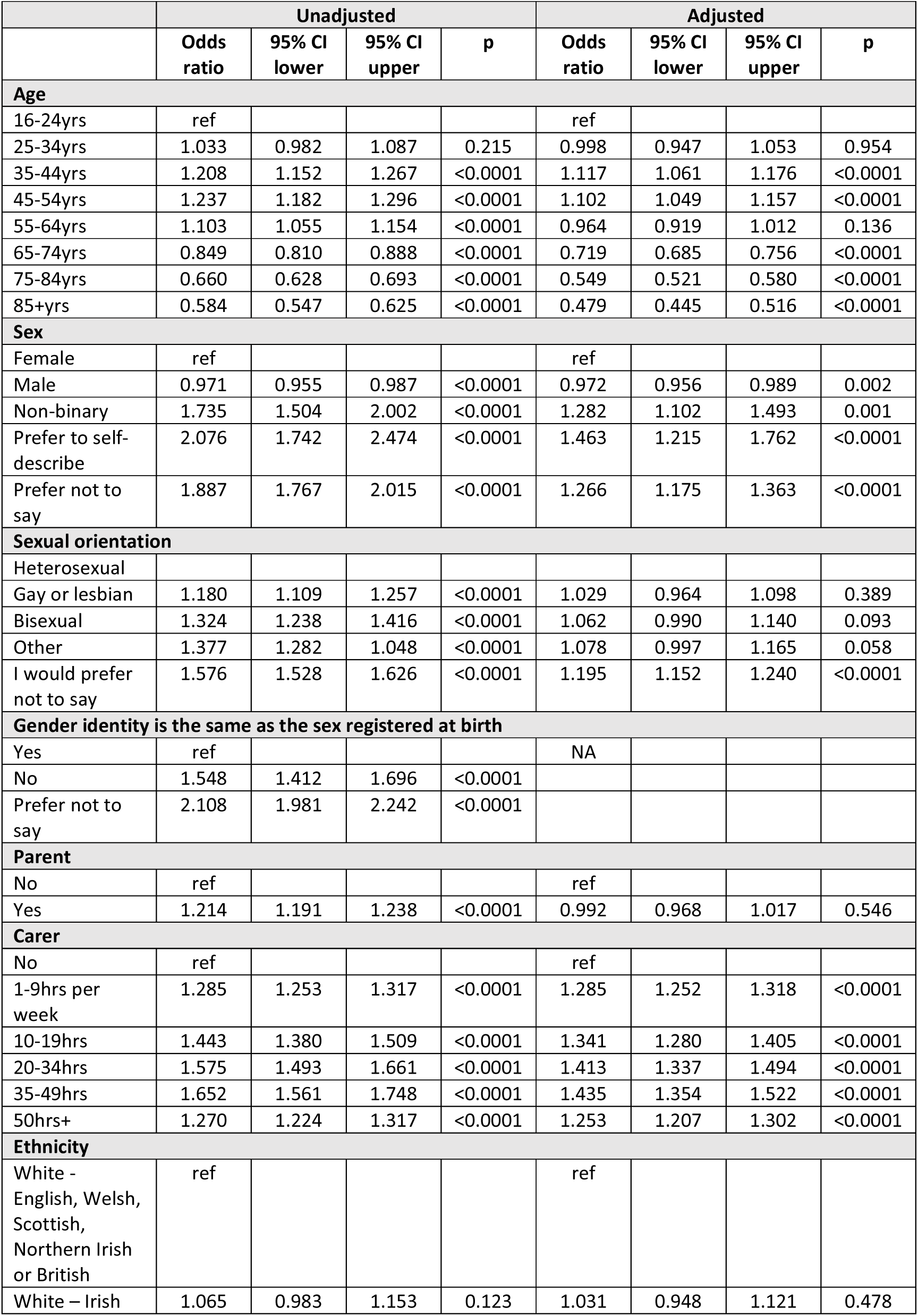
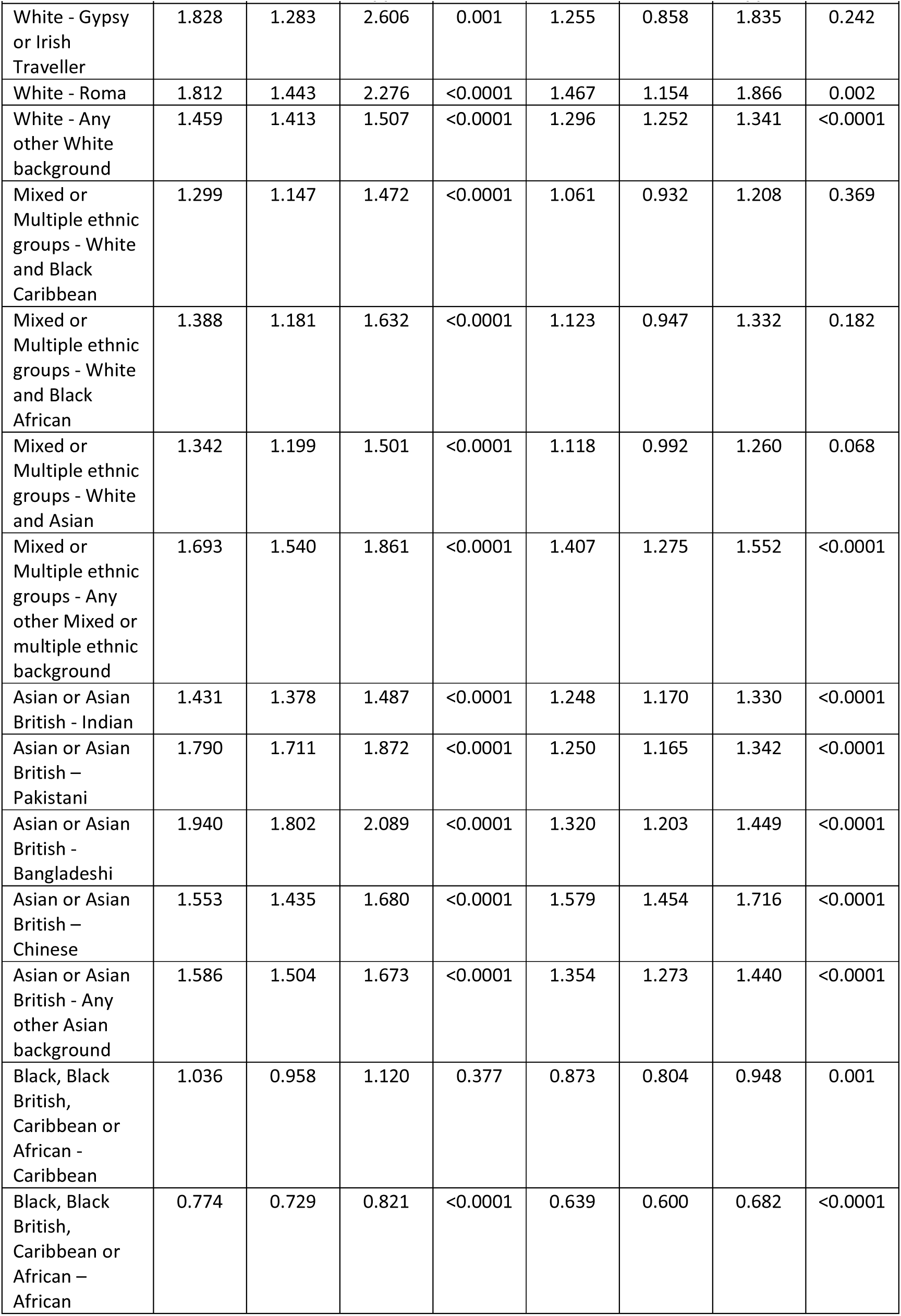
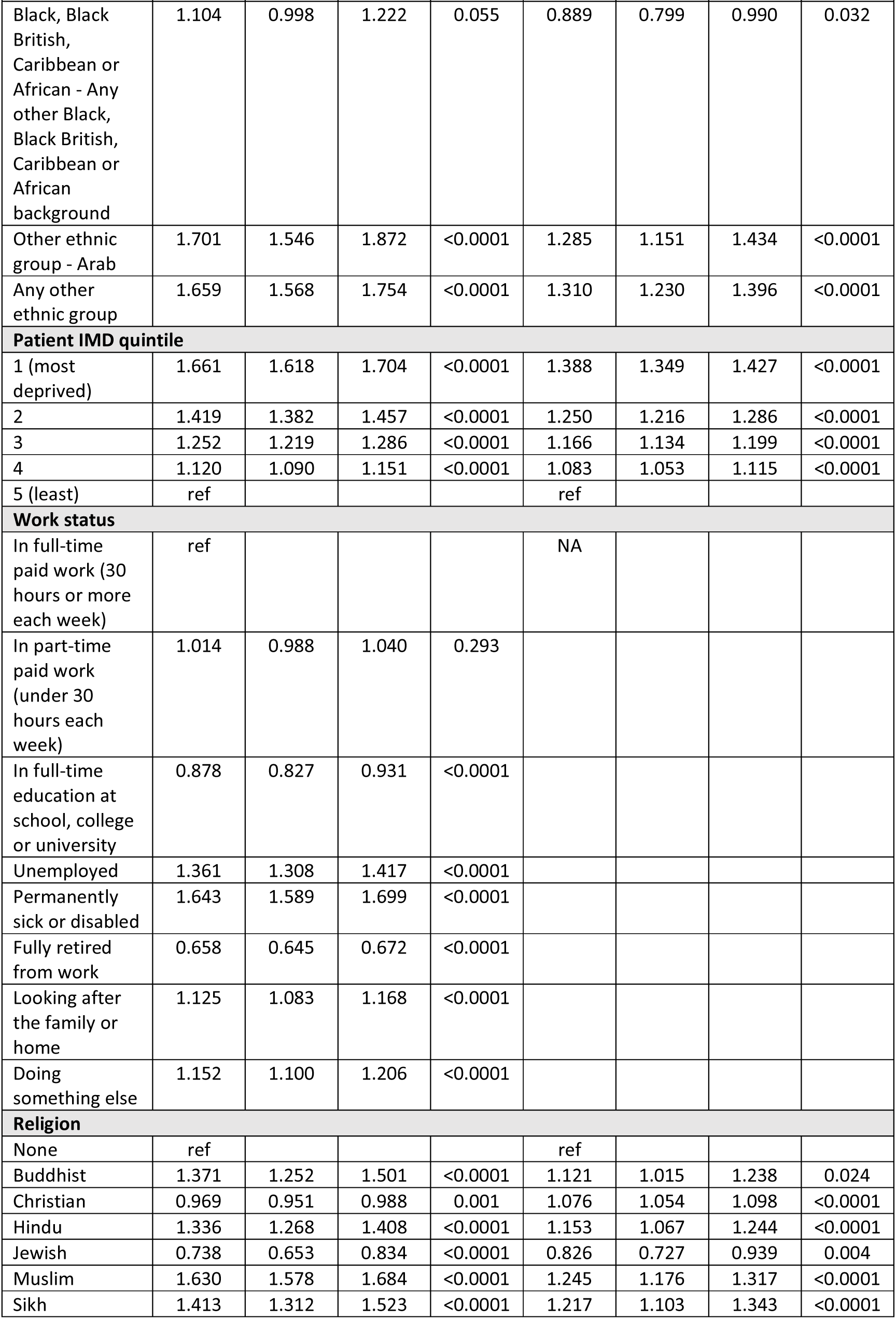
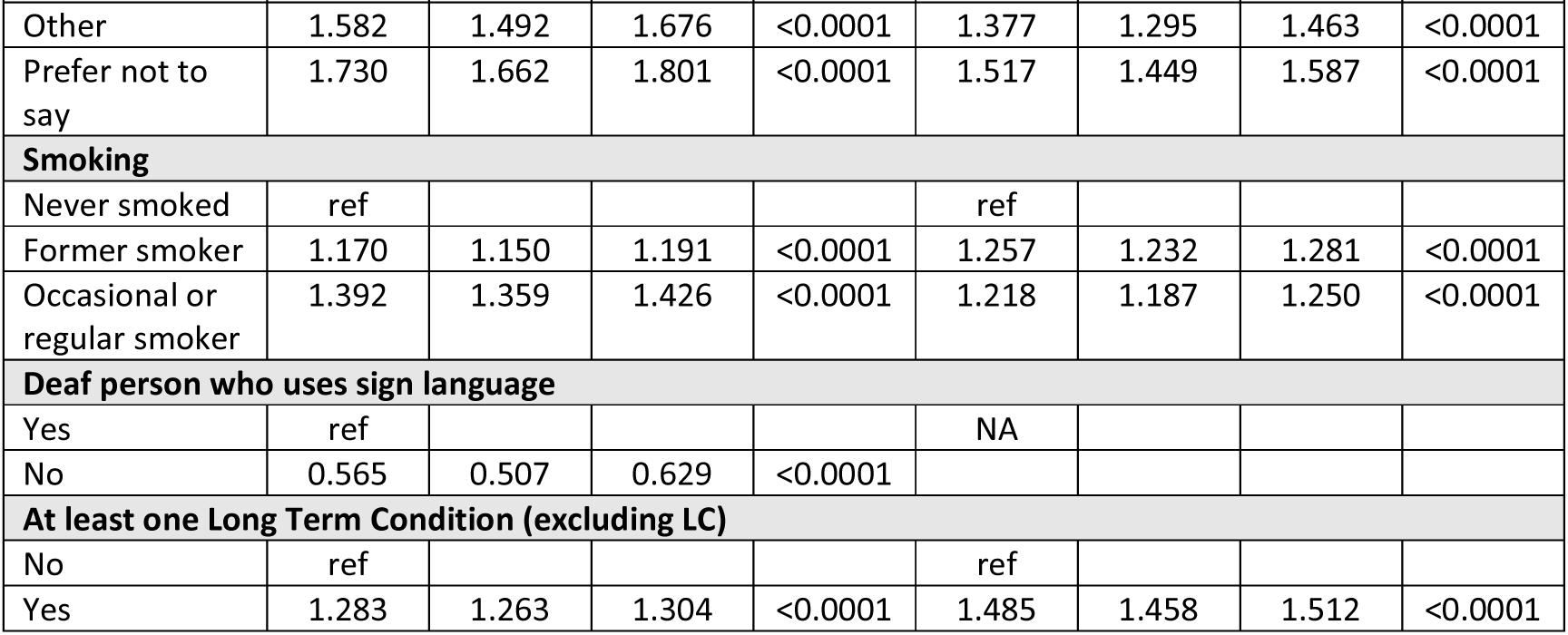
Odds of being uncertain about having Long Covid compared to not having Long Covid (not sure vs no) using General Practice Patient Survey data (2023)

|  | Unadjusted |  |  |  | Adjusted |  |  |  |
| --- | --- | --- | --- | --- | --- | --- | --- | --- |
|  | Odds ratio | 95% CI lower | 95% CI upper | p | Odds ratio | 95% CI lower | 95% CI upper | p |
| **Age** |  |  |  |  |  |  |  |  |
| 16-24yrs | ref |  |  |  | ref |  |  |  |
| 25-34yrs | 1.033 | 0.982 | 1.087 | 0.215 | 0.998 | 0.947 | 1.053 | 0.954 |
| 35-44yrs | 1.208 | 1.152 | 1.267 | <0.0001 | 1.117 | 1.061 | 1.176 | <0.0001 |
| 45-54yrs | 1.237 | 1.182 | 1.296 | <0.0001 | 1.102 | 1.049 | 1.157 | <0.0001 |
| 55-64yrs | 1.103 | 1.055 | 1.154 | <0.0001 | 0.964 | 0.919 | 1.012 | 0.136 |
| 65-74yrs | 0.849 | 0.810 | 0.888 | <0.0001 | 0.719 | 0.685 | 0.756 | <0.0001 |
| 75-84yrs | 0.660 | 0.628 | 0.693 | <0.0001 | 0.549 | 0.521 | 0.580 | <0.0001 |
| 85+yrs | 0.584 | 0.547 | 0.625 | <0.0001 | 0.479 | 0.445 | 0.516 | <0.0001 |
| **Sex** |  |  |  |  |  |  |  |  |
| Female | ref |  |  |  | ref |  |  |  |
| Male | 0.971 | 0.955 | 0.987 | <0.0001 | 0.972 | 0.956 | 0.989 | 0.002 |
| Non-binary | 1.735 | 1.504 | 2.002 | <0.0001 | 1.282 | 1.102 | 1.493 | 0.001 |
| Prefer to self-describe | 2.076 | 1.742 | 2.474 | <0.0001 | 1.463 | 1.215 | 1.762 | <0.0001 |
| Prefer not to say | 1.887 | 1.767 | 2.015 | <0.0001 | 1.266 | 1.175 | 1.363 | <0.0001 |
| **Sexual orientation** |  |  |  |  |  |  |  |  |
| Heterosexual |  |  |  |  |  |  |  |  |
| Gay or lesbian | 1.180 | 1.109 | 1.257 | <0.0001 | 1.029 | 0.964 | 1.098 | 0.389 |
| Bisexual | 1.324 | 1.238 | 1.416 | <0.0001 | 1.062 | 0.990 | 1.140 | 0.093 |
| Other | 1.377 | 1.282 | 1.048 | <0.0001 | 1.078 | 0.997 | 1.165 | 0.058 |
| I would prefer not to say | 1.576 | 1.528 | 1.626 | <0.0001 | 1.195 | 1.152 | 1.240 | <0.0001 |
| **Gender identity is the same as the sex registered at birth** |  |  |  |  |  |  |  |  |
| Yes | ref |  |  |  | NA |  |  |  |
| No | 1.548 | 1.412 | 1.696 | <0.0001 |  |  |  |  |
| Prefer not to say | 2.108 | 1.981 | 2.242 | <0.0001 |  |  |  |  |
| **Parent** |  |  |  |  |  |  |  |  |
| No | ref |  |  |  | ref |  |  |  |
| Yes | 1.214 | 1.191 | 1.238 | <0.0001 | 0.992 | 0.968 | 1.017 | 0.546 |
| **Carer** |  |  |  |  |  |  |  |  |
| No | ref |  |  |  | ref |  |  |  |
| 1-9hrs per week | 1.285 | 1.253 | 1.317 | <0.0001 | 1.285 | 1.252 | 1.318 | <0.0001 |
| 10-19hrs | 1.443 | 1.380 | 1.509 | <0.0001 | 1.341 | 1.280 | 1.405 | <0.0001 |
| 20-34hrs | 1.575 | 1.493 | 1.661 | <0.0001 | 1.413 | 1.337 | 1.494 | <0.0001 |
| 35-49hrs | 1.652 | 1.561 | 1.748 | <0.0001 | 1.435 | 1.354 | 1.522 | <0.0001 |
| 50hrs+ | 1.270 | 1.224 | 1.317 | <0.0001 | 1.253 | 1.207 | 1.302 | <0.0001 |
| **Ethnicity** |  |  |  |  |  |  |  |  |
| White - English, Welsh, Scottish, Northern Irish or British | ref |  |  |  | ref |  |  |  |
| White – Irish | 1.065 | 0.983 | 1.153 | 0.123 | 1.031 | 0.948 | 1.121 | 0.478 |
|  | Odds ratio | 95% CI lower | 95% CI upper | p | Odds ratio | 95% CI lower | 95% CI upper | p |
| White - Gypsy or Irish Traveller | 1.828 | 1.283 | 2.606 | 0.001 | 1.255 | 0.858 | 1.835 | 0.242 |
| White - Roma | 1.812 | 1.443 | 2.276 | <0.0001 | 1.467 | 1.154 | 1.866 | 0.002 |
| White - Any other White background | 1.459 | 1.413 | 1.507 | <0.0001 | 1.296 | 1.252 | 1.341 | <0.0001 |
| Mixed or Multiple ethnic groups - White and Black Caribbean | 1.299 | 1.147 | 1.472 | <0.0001 | 1.061 | 0.932 | 1.208 | 0.369 |
| Mixed or Multiple ethnic groups - White and Black African | 1.388 | 1.181 | 1.632 | <0.0001 | 1.123 | 0.947 | 1.332 | 0.182 |
| Mixed or Multiple ethnic groups - White and Asian | 1.342 | 1.199 | 1.501 | <0.0001 | 1.118 | 0.992 | 1.260 | 0.068 |
| Mixed or Multiple ethnic groups - Any other Mixed or multiple ethnic background | 1.693 | 1.540 | 1.861 | <0.0001 | 1.407 | 1.275 | 1.552 | <0.0001 |
| Asian or Asian British - Indian | 1.431 | 1.378 | 1.487 | <0.0001 | 1.248 | 1.170 | 1.330 | <0.0001 |
| Asian or Asian British – Pakistani | 1.790 | 1.711 | 1.872 | <0.0001 | 1.250 | 1.165 | 1.342 | <0.0001 |
| Asian or Asian British - Bangladeshi | 1.940 | 1.802 | 2.089 | <0.0001 | 1.320 | 1.203 | 1.449 | <0.0001 |
| Asian or Asian British – Chinese | 1.553 | 1.435 | 1.680 | <0.0001 | 1.579 | 1.454 | 1.716 | <0.0001 |
| Asian or Asian British - Any other Asian background | 1.586 | 1.504 | 1.673 | <0.0001 | 1.354 | 1.273 | 1.440 | <0.0001 |
| Black, Black British, Caribbean or African - Caribbean | 1.036 | 0.958 | 1.120 | 0.377 | 0.873 | 0.804 | 0.948 | 0.001 |
| Black, Black British, Caribbean or African – African | 0.774 | 0.729 | 0.821 | <0.0001 | 0.639 | 0.600 | 0.682 | <0.0001 |
|  | Odds ratio | 95% CI lower | 95% CI upper | p | Odds ratio | 95% CI lower | 95% CI upper | p |
| Black, Black British, Caribbean or African - Any other Black, Black British, Caribbean or African background | 1.104 | 0.998 | 1.222 | 0.055 | 0.889 | 0.799 | 0.990 | 0.032 |
| Other ethnic group - Arab | 1.701 | 1.546 | 1.872 | <0.0001 | 1.285 | 1.151 | 1.434 | <0.0001 |
| Any other ethnic group | 1.659 | 1.568 | 1.754 | <0.0001 | 1.310 | 1.230 | 1.396 | <0.0001 |
| **Patient IMD quintile** |  |  |  |  |  |  |  |  |
| 1 (most deprived) | 1.661 | 1.618 | 1.704 | <0.0001 | 1.388 | 1.349 | 1.427 | <0.0001 |
| 2 | 1.419 | 1.382 | 1.457 | <0.0001 | 1.250 | 1.216 | 1.286 | <0.0001 |
| 3 | 1.252 | 1.219 | 1.286 | <0.0001 | 1.166 | 1.134 | 1.199 | <0.0001 |
| 4 | 1.120 | 1.090 | 1.151 | <0.0001 | 1.083 | 1.053 | 1.115 | <0.0001 |
| 5 (least) | ref |  |  |  | ref |  |  |  |
| **Work status** |  |  |  |  |  |  |  |  |
| In full-time paid work (30 hours or more each week) | ref |  |  |  | NA |  |  |  |
| In part-time paid work (under 30 hours each week) | 1.014 | 0.988 | 1.040 | 0.293 |  |  |  |  |
| In full-time education at school, college or university | 0.878 | 0.827 | 0.931 | <0.0001 |  |  |  |  |
| Unemployed | 1.361 | 1.308 | 1.417 | <0.0001 |  |  |  |  |
| Permanently sick or disabled | 1.643 | 1.589 | 1.699 | <0.0001 |  |  |  |  |
| Fully retired from work | 0.658 | 0.645 | 0.672 | <0.0001 |  |  |  |  |
| Looking after the family or home | 1.125 | 1.083 | 1.168 | <0.0001 |  |  |  |  |
| Doing something else | 1.152 | 1.100 | 1.206 | <0.0001 |  |  |  |  |
| **Religion** |  |  |  |  |  |  |  |  |
| None | ref |  |  |  | ref |  |  |  |
| Buddhist | 1.371 | 1.252 | 1.501 | <0.0001 | 1.121 | 1.015 | 1.238 | 0.024 |
| Christian | 0.969 | 0.951 | 0.988 | 0.001 | 1.076 | 1.054 | 1.098 | <0.0001 |
| Hindu | 1.336 | 1.268 | 1.408 | <0.0001 | 1.153 | 1.067 | 1.244 | <0.0001 |
| Jewish | 0.738 | 0.653 | 0.834 | <0.0001 | 0.826 | 0.727 | 0.939 | 0.004 |
| Muslim | 1.630 | 1.578 | 1.684 | <0.0001 | 1.245 | 1.176 | 1.317 | <0.0001 |
| Sikh | 1.413 | 1.312 | 1.523 | <0.0001 | 1.217 | 1.103 | 1.343 | <0.0001 |
|  | Odds ratio | 95% CI lower | 95% CI upper | p | Odds ratio | 95% CI lower | 95% CI upper | p |
| Other | 1.582 | 1.492 | 1.676 | <0.0001 | 1.377 | 1.295 | 1.463 | <0.0001 |
| Prefer not to say | 1.730 | 1.662 | 1.801 | <0.0001 | 1.517 | 1.449 | 1.587 | <0.0001 |
| **Smoking** |  |  |  |  |  |  |  |  |
| Never smoked | ref |  |  |  | ref |  |  |  |
| Former smoker | 1.170 | 1.150 | 1.191 | <0.0001 | 1.257 | 1.232 | 1.281 | <0.0001 |
| Occasional or regular smoker | 1.392 | 1.359 | 1.426 | <0.0001 | 1.218 | 1.187 | 1.250 | <0.0001 |
| **Deaf person who uses sign language** |  |  |  |  |  |  |  |  |
| Yes | ref |  |  |  | NA |  |  |  |
| No | 0.565 | 0.507 | 0.629 | <0.0001 |  |  |  |  |
| **At least one Long Term Condition (excluding LC)** |  |  |  |  |  |  |  |  |
| No | ref |  |  |  | ref |  |  |  |
| Yes | 1.283 | 1.263 | 1.304 | <0.0001 | 1.485 | 1.458 | 1.512 | <0.0001 |

## Discussion

Our analysis of this national general practice survey data found that 4.8% of respondents self-described as having LC while 9.1% were unsure whether they have it or not. There is an unequal distribution of LC in England, with the condition being more prevalent in minoritised and disadvantaged groups. There are also high levels of uncertainty about LC, with certain groups that are already disadvantaged being more likely to be uncertain if they have the condition.

Many of this study’s findings support existing research on risk factors for LC, for example lower odds of having LC in males compared to females^2–9^, higher odds in those of working age^2–4, 6, 7^, the strong trend of increasing odds of LC with increasing deprivation^1, 3–5, 9^ and higher odds for people who are unemployed^1^, and who have existing health conditions^2, 5, 7^. UK studies have also found higher prevalence of LC in particular ethnic minority groups^1, 3, 5, 8^ but findings have not been consistent. Lower odds were also found for people of Black ethnicity, something that has been found in other UK studies^1, 7^. Higher prevalence in people from sexual orientation minorities has also been identified in one US study^23^.

In this study, there were higher odds of having LC for people who are parents and carers. This could be explained by disproportionate exposure to SARS-CoV-2 through schools and caring roles, or greater awareness of LC in these groups. The latter is supported by the reduced odds found in this study of being unsure about having LC in those groups. The impact of LC symptoms could be more noticeable due to caring roles, or the likelihood of being able to rest during the acute COVID-19 phase may have been limited (a potential factor in some LC cases^10^). There were also higher odds for people of particular faiths, even after adjusting for ethnicity, deprivation, age, gender and other potential confounding factors. However, this still may be explained by residual confounding. Odds of having LC were higher in former smokers but lower for current smokers, which is contrary to studies where smoking has been identified as a risk factor^3, 5^. However, reviews of this area recognise the variation in findings about the role of smoking in LC^24^.

This study also adds new evidence about the lack of certainty people feel about whether or not they have LC. Groups who were less confident about whether they had LC included males and non-binary people, people from ‘Any other white background’, Indian, Bangladeshi, Chinese, ‘Any other Asian background’, Black Caribbean, Black African, Arab and ‘Any other ethnic group’, those who were working part-time, unemployed, retired, or staying at home and smokers. Those who were less likely to be unsure about having LC included those aged 24-74yrs, those who were not heterosexual, parents/guardians and carers, White Gypsy/Irish Traveller, people who were sick or disabled and people with at least one defined LTC. Those under the age of 25yrs were less confident about having LC. This could explain why most research about LC includes older people as most research uses self-identification with LC as study inclusion strategy. It could also be the case that there is wider associated stigma with having LC in younger people^17^.

Given the high burden of LC in the global population and its impact on everyday lives of individuals and the functioning of society, it is important to understand the drivers for uncertainty around LC to enable people to get the right support. Levels of uncertainty may indicate a lack of awareness of LC symptoms or confusion, or there may be an overlap in symptoms with other conditions. A lack of uncertainty in particular groups may be because a diagnosis has already been secured, indicating that some groups may be more likely than others to be successfully diagnosed, or it could be related to levels of health literacy and self-advocacy. Stigma and self-doubt can shape self-identification with LC^16, 17, 25, 26^ which could be discouraging people from coming forward for diagnosis and support and there is emerging evidence that intersectional identity may compound LC stigma leading to worse outcomes for marginalised groups^16^.

### Strengths and Limitations

A key strength of this study is its large sample size and a national survey sample. Limitations of this study include the lack of data about severity or symptoms of LC, the timing of its onset and the COVID-19 vaccination status of respondents. In addition, interpreting survey responses about LTCs with respect to LC is problematic. LC is a long-term condition but 21.9% of people who described themselves as having LC said they did not have any LTCs. To take account of this the multivariable models in this study included a variable constructed from named LTCs as reported by respondents rather than the answer to the question ‘any other LTC’ to eliminate potentially including the LC outcome. A further limitation is that we cannot role reverse causality for some of the variables such as work status, hence this variable was not included in the multivariable analysis.

### Conclusions

This study provides further evidence that particular population groups in England, often those minoritised and disadvantaged, are more likely to have LC. It adds new evidence that there are more people who are unsure whether they have LC than those who are confident they have it, again some from more vulnerable groups. This clearly demonstrable health inequality lends weight to the argument for diagnosis, treatment and support to be better distributed according to need^27^, to avoid LC becoming another factor contributing to the widening health gap between different groups in society. Improved awareness about the condition amongst the general population and training about barriers and stigma among healthcare professionals are required to ensure people with LC can be identified and provided with the right treatment and support.

## Data Availability

GP Patient Survey data is available at GP practice-level via https://gp-patient.co.uk/analysistool. Requests for GPPS patient-level datasets can be made directly to the survey team whose details can be found here https://gp-patient.co.uk/confidentiality

## References

1. Office for National Statistics. Self-reported coronavirus (COVID-19) infections and associated symptoms, England and Scotland: November 2023 to March 2024. 25 April 2024 2024. Online: Office for National Statistics.

2. Luo D, Mei B, Wang P, et al. Prevalence and risk factors for persistent symptoms after COVID-19: a systematic review and meta-analysis. Clinical Microbiology and Infection 2024; 30: 328–335. DOI: 10.1016/j.cmi.2023.10.016.

3. Whitaker M, Elliott J, Chadeau-Hyam M, et al. Persistent COVID-19 symptoms in a community study of 606,434 people in England. Nature Communications 2022; 13: 1957. DOI: 10.1038/s41467-022-29521-z.

4. Hastie CE, Lowe DJ, McAuley A, et al. Outcomes among confirmed cases and a matched comparison group in the Long-COVID in Scotland study. Nature Communications 2022; 13: 5663. DOI: 10.1038/s41467-022-33415-5.

5. Subramanian A, Nirantharakumar K, Hughes S, et al. Symptoms and risk factors for long COVID in non-hospitalized adults. Nature Medicine 2022; 28: 1706–1714. DOI: 10.1038/s41591-022-01909-w.

6. Evans RA, McAuley H, Harrison EM, et al. Physical, cognitive, and mental health impacts of COVID-19 after hospitalisation (PHOSP-COVID): a UK multicentre, prospective cohort study. The Lancet Respiratory Medicine 2021; 9: 1275–1287. DOI: 10.1016/S2213-2600(21)00383-0.

7. Thompson EJ, Williams DM, Walker AJ, et al. Long COVID burden and risk factors in 10 UK longitudinal studies and electronic health records. Nature Communications 2022; 13: 3528. DOI: 10.1038/s41467-022-30836-0.

8. Wang HI, Doran T, Crooks M, et al. EPH11 Prevalence, Risk Factors, and Characterization of Individuals with Long COVID Using Electronic Health Records in Over 1.5 Million COVID Cases in England. Value in Health 2023; 26: S204. DOI: 10.1016/j.jval.2023.09.1053.

9. Shabnam S, Razieh C, Dambha-Miller H, et al. Socioeconomic inequalities of Long COVID: a retrospective population-based cohort study in the United Kingdom. Journal of the Royal Society of Medicine 2023; 116: 263–273. DOI: 10.1177/01410768231168377.

10. Ziauddeen N, Gurdasani D, O’Hara ME, et al. Characteristics and impact of Long Covid: Findings from an online survey. PLOS ONE 2022; 17: e0264331. DOI: 10.1371/journal.pone.0264331.

11. Davis HE, Assaf GS, McCorkell L, et al. Characterizing long COVID in an international cohort: 7 months of symptoms and their impact. eClinicalMedicine 2021; 38. DOI: 10.1016/j.eclinm.2021.101019.

12. Office for National Statistics. Coronavirus and the social impacts of ‘long COVID’ on people’s lives in Great Britain: 7 April to 13 June 2021. 2021. Online.

13. Baz SA, Fang C, Carpentieri JD and Sheard L. ‘I don’t know what to do or where to go’. Experiences of accessing healthcare support from the perspectives of people living with Long Covid and healthcare professionals: A qualitative study in Bradford, UK. Health Expectations 2023; 26: 542–554. 10.1111/hex.13687. DOI: 10.1111/hex.13687.

14. Smyth N, Gaszczyk P, Alwan NA, et al. OP63 ‘You can be in hell and they still refuse to help’: Racially and ethnically minoritised people with Long COVID reflect on poor care and support experiences. Journal of Epidemiology and Community Health 2023; 77: A31. DOI: 10.1136/jech-2023-SSMabstracts.62.

15. Dean E. What happens inside a long covid clinic? BMJ 2023; 382: p 1791. DOI: 10.1136/bmj.p1791.

16. Clutterbuck D, Ramasawmy M, Pantelic M, et al. Barriers to healthcare access and experiences of stigma: Findings from a coproduced Long Covid case-finding study. Health Expectations 2024; 27: e14037. DOI: 10.1111/hex.14037.

17. Pantelic M, Ziauddeen N, Boyes M, et al. Long Covid stigma: Estimating burden and validating scale in a UK-based sample. PLOS ONE 2022; 17: e0277317. DOI: 10.1371/journal.pone.0277317.

18. NHS England Ipsos. GP Patient Survey. Online 2022.

19. NHS England Ipsos. GP Patient Survey 2023 Technical Annex. 2023.

20. NHS England Ipsos. GP Patient Survey questionnaire. 2023.

21. StataCorp. Stata Statistical Software: Release 17. College Station, TX: StataCorp LLC, 2021.

22. Noble S, McLennan D, Noble M, et al. The English Indices of Deprivation 2019: Research report. In: Ministry of Housing, Communities and Local Government, (ed.). 2019.

23. Cohen J and van der Meulen Rodgers Y. An intersectional analysis of long COVID prevalence. International Journal for Equity in Health 2023; 22: 261. DOI: 10.1186/s12939-023-02072-5.

24. Trofor AC, Robu Popa D, Melinte OE, et al. Looking at the Data on Smoking and Post-COVID-19 Syndrome—A Literature Review. Journal of Personalized Medicine 14. DOI: 10.3390/jpm14010097.

25. Smyth N, Ridge D, Kingstone T, et al. People from ethnic minorities seeking help for Long Covid: a qualitative study. British Journal of General Practice 2024: BJGP.2023.0631. DOI: 10.3399/BJGP.2023.0631.

26. Ireson J, Taylor A, Richardson E, et al. Exploring invisibility and epistemic injustice in Long Covid—A citizen science qualitative analysis of patient stories from an online Covid community. Health Expectations 2022; 25: 1753–1765. 10.1111/hex.13518.

27. Hutchinson J, Checkland K, Munford L, et al. Long COVID in general practice: an analysis of the equity of NHS England’s enhanced service specification. British Journal of General Practice 2022; 72: 85. DOI: 10.3399/bjgp22X718505.

